# Validation of the *Mayo-Portland Adaptability Inventory-4 (MPAI-4)* and reference norms in a French-Canadian population with traumatic brain injury receiving rehabilitation

**DOI:** 10.1101/2020.10.26.20219956

**Authors:** Marie-Claude Guerrette, Michelle McKerral

## Abstract

**Objective:** To validate the factor structure and establish internal consistency reliability of the French-Canadian version of the Mayo-Portland Adaptability Inventory (MPAI-4), using a Canadian sample of adults with traumatic brain injury (TBI) receiving post-acute rehabilitation services.

**Design:** Psychometric analysis of prospectively collected French-Canadian MPAI-4 data.

**Setting:** Inpatient and outpatient TBI rehabilitation programs.

**Participants:** Adults (*N* = 1012) with a mild, moderate or severe TBI who received inpatient or outpatient rehabilitation interventions and for whom a first French-Canadian MPAI-4 measure was completed between 2016 and 2020.

**Interventions:** Not applicable.

**Main Outcome Measure:** French-Canadian MPAI-4 questionnaire.

**Results:** To evaluate the factor structure of the French-Canadian MPAI-4, an exploratory factor analysis using a varimax rotation method was conducted on z-scores for all items. The final and best solution was a three-factor solution, which accounted for 48.68% of the variance. The internal consistency of the French-Canadian MPAI-4 was determined using Cronbach’s alpha and all three subscales showed good internal consistency (all .70 *≤ α ≤* .89). Reference norms for the TBI sample are provided in the form of T scores for subscales and total score, as well as descriptive raw data according to sex, age, TBI severity and rehabilitation setting.

**Conclusions:** The three factors extracted using data from the French-Canadian MPAI-4 are similar, but not entirely identical to the three subscales of the original MPAI-4. Overall, the French-Canadian MPAI-4 factor structure is validated. The questionnaire shows good psychometric properties and represents a suitable tool to measure functional evolution and social participation of TBI adults receiving rehabilitation services in a French-Canadian context. The provided reference norms will also help guide the clinical use of the MPAI-4 in French-Canadian TBI populations.

---

Traumatic brain injury (TBI) is the leading cause of death and disability across the world and consists of a serious health problem.^1^ It also remains a complex medical condition because patients are at high risk of having long-lasting physical, cognitive or behavioural sequelae that may impact their social integration or vocational recovery. Therefore, the use of standardized, valid and reliable tools that measures sequelae and help professionals identify functional impairment and predict the outcome is necessary,^2^ such as during the assessment following TBI. However, considering the complexity and heterogeneity of TBI sequelae, there are few available tools that measure TBI impairments and participation levels, as most of the accessible measurement tools are generic and not specific to TBI.^3–5^

One measurement tool commonly used by rehabilitation professionals working with TBI population is the Mayo-Portland Adaptability Inventory-4 questionnaire (MPAI-4),^6^ developed in the United States of America. Lezak^7^ produced the initial Portland Adaptability Inventory (PAI), which was refined by Malec and Thompson^8^ and renamed the Mayo-Portland Adaptability Inventory (MPAI). Over the years, the MPAI was further modified to maximize its internal consistency and reliability (for example, see Malec et al.^9^), resulting in its most recent version, the Mayo-Portland Adaptability Inventory, 4^th^ revision (MPAI-4).^6^ The MPAI-4 questionnaire was broadly designed to assess functional abilities, global outcome and community integration by covering a wide range of physical, cognitive, emotional, behavioural and social problems that TBI patients may experience following their brain injury.

The MPAI-4 is a 30-item questionnaire measuring the patient’s magnitude of impairments. The 30 items are divided into three subscales (i.e. Ability, Adjustment, Participation), with some overlap in categories. The Ability index consists of 13 items measuring mobility, hand function, vision, hearing, dizziness, motor speech, verbal and nonverbal communication, attention and concentration, memory, information retrieval, problem solving and visuospatial abilities. The Adjustment index, with 12 items, assesses anxiety, depression, irritability and anger or aggression, pain and headaches, fatigue, sensitivity to mild symptoms, inappropriate social interactions, impaired self-awareness, family and significant relationships, initiative, social contact, and leisure and recreational activities. The Participation index, with 8 items, measures initiative, social contact, leisure and recreational activities (i.e. the latter three items contribute to both the Adjustment index and the Participation index), self-care, independent living, independent use of transportation, employment status and financial management. Each item is scored on a 5-point scale (from 0 to 4), with higher score indicating greater clinical impairment. Each subscale evaluates different aspects of TBI sequelae, and the subscales can be used separately or combined in a total MPAI-4 score, reflecting the general level of adaptation and social participation.

The MPAI-4 questionnaire is now extensively used worldwide in inpatient, outpatient and vocational rehabilitation settings to measure TBI patients’ progress and outcomes.^10–14^ The multifaceted structure of the MPAI-4 makes it a useful tool for planning and assessing interventions, and studies have demonstrated the MPAI-4’s clinical sensitivity to the effect of rehabilitation.^15–17^ The MPAI-4 is best completed by consensus of the rehabilitation team, but can be completed by caregivers, significant others or by the patients themselves. For example, Malec^18^ compared MPAI-4 scores between rehabilitation professionals, significant others and TBI patients, which revealed satisfactory internal consistency and interrater agreement. Finally, the MPAI-4 is among the recommended Common Data Elements Project for TBI adults, and an emerging measure for youth with TBI.^19,20^ It is also a recommended participation measure in the INESSS-ONF Clinical Practice Guidelines following moderate and severe TBI.^21^

Using Rasch analysis, item clusters, principal component analyses and other traditional psychometric measures, numerous studies provided further evidence of satisfactory internal consistency,^9,16,22^ construct validity,^9,16,17,23^ as well as predictive and concurrent validity^8,23–28^ for the full MPAI-4 measure and its subscales. Moreover, the MPAI-4 also proved itself to be a useful measure of global outcome for patients following a stroke.^29,30^

Overall, given its clinical usefulness, comprehensiveness and good psychometric properties, the MPAI-4 questionnaire was translated into other languages, notably French, Danish, Spanish, German, Italian, Portuguese, Swedish and Dutch. However, aside from the original MPAI-4, psychometric properties have only been established for the Italian^4^ and Arabic^31^ versions of the MPAI-4. Thus, the objective of this paper was to validate the factor structure and establish internal consistency reliability of the French-Canadian MPAI-4, using a Canadian sample of TBI adults receiving post-acute rehabilitation services in a French-speaking environment. A secondary aim was to provide a set of French-Canadian MPAI-4 reference norms for TBI to guide clinical use.

## Methods

### French-Canadian version of the Mayo-Portland Adaptability Inventory-4

With the permission and collaboration of the MPAI-4’s author (James F. Malec), the questionnaire was translated into French and adapted to a French-Canadian cultural context. The original MPAI-4 questionnaire was translated into French by a professional scientific translator and was then back-translated into English. The two versions were then submitted to an expert multidisciplinary clinical team for review (neuropsychologist, occupational therapist, physiotherapist, social worker) in order to choose the most appropriate terms in French for describing the different MPAI-4 items. The resulting version was then submitted to a second panel of clinicians (neuropsychologist, occupational therapist) for final validation of the French-Canadian questionnaire’s terminology. The resulting French-Canadian version of the MPAI-4 and its user’s manual can be found on the COMBI website.^32–34^

### Procedure and Participants

Between 2014 and 2017, the French-Canadian MPAI-4 was implemented in the clinical practice of four rehabilitation centres in the greater Montreal region. Since 2016, for every TBI patient receiving post-acute rehabilitation services, the MPAI-4 is completed by team consensus at the beginning of inpatient rehabilitation, and at the beginning and end of outpatient rehabilitation. For the purpose of this study, all patients’ first MPAI-4 measure (at intake either for inpatient or outpatient rehabilitation) was used in the analysis. Eligible patients (*N* = 1020) were TBI adults participating in a TBI rehabilitation program at one of the four rehabilitation centres, and for whom at least a first MPAI-4 measure was completed between 2016 and 2020. Data from 1012 of these patients were included in the final analyses. The patients’ sociodemographic and clinical characteristics are presented in Table 1. Ethical approval for collecting patients’ MPAI-4 scores, sociodemographic and clinical characteristics was obtained from each rehabilitation centre’s research ethics board.

**Table 1.** Patients’ sociodemographic and clinical characteristics.

|  | Total *N* = 1012 (%) |
| --- | --- |
| Mean age (in years) at TBI (*N* = 1012) | 55.83 ± 21.07 |
| Sex |  |
| Male | 625 (61.80) |
| Female | 386 (38.10) |
| Missing | 1 (0.10) |
| TBI severity |  |
| Mild | 229 (22.60) |
| Complicated mild | 229 (22.60) |
| Moderate | 371 (36.70) |
| Severe | 183 (18.10) |
| Education level |  |
| University | 284 (28.10) |
| College | 221 (21.80) |
| High school | 223 (22.00) |
| Elementary | 216 (21.30) |
| None | 38 (3.80) |
| Missing | 30 (3.00) |
| Mean time (in months) between TBI and MP AI-4 measure (*N* = 1011) | 4.93 ± 14.16 |

### Statistical Analysis

An exploratory factor analysis (EFA) using orthogonal (varimax) rotation method was used to evaluate the construct validity of the French-Canadian MPAI-4. The Kaiser-Meyer-Olkin (KMO) measure verified the sampling adequacy for the EFA and the Bartlett’s test of sphericity was used to assess the degree of inter-correlation between variables. Analyzed matrices (correlation, anti-image and reproduced correlations) are available upon request. Factors were selected if the eigenvalue was > 1 as suggested by Kaiser^35^ and the number of factors was validated by inspection of the scree plot^36,37^ with clinical consideration for coherence and interpretability of the factors. Appropriate for our sample size, conservative cutoffs of .20 on loading values were applied for statistical significance of an item^38^ and items with loading values *≥* .40 were included in the interpretation of a factor.^36,37^ The internal consistency of the French-Canadian MPAI-4 was determined using Cronbach’s alpha. For all analyses, the level of significance was .05 using 95% confidence intervals. Finally, French-Canadian reference norms were developed for the MPAI-4 total score and subscales by converting raw scores into standardized T scores (Mean = 50; SD = 10). Means and SD were also computed on raw MPAI-4 data according to sex, age, TBI severity and rehabilitation setting. Analyses were conducted using IBM SPSS Statistics for Macintosh version 26.0 (2019).

## Results

Prior to the factor analysis, raw MPAI-4 scores were examined for accuracy of data entry, missing values, outliers and fit between data distribution and the assumptions of multivariate analyses. The variables were examined separately for all participants. No missing values were found. No extremely low or high z-scores were found to be univariate outliers, but eight cases were identified as multivariate outliers using Mahalanobis distance (*p* < .001). The eight participants were removed. The final sample size of 1012 participants is adequate for factor analysis according to literature guidelines.^39^

### Exploratory factor analysis (EFA)

Before conducting the factor analysis on the full sample, participants were randomly divided into two groups. Exploratory factor analyses (EFA) with varimax rotation methods were performed separately on z-scores from both groups. The number of extracted factors, factor loadings for individual items as well as proportion of variance explained by each factor was similar between groups. The random samples were therefore combined and the EFA was performed on the whole sample. The EFA with varimax rotation method was performed on z-scores for the 30 items of the French-Canadian MPAI-4. The overall KMO was .93 and all KMO value for individual items were larger than or equal to .76, which is above the acceptable limit of 50.36 Bartlett’s test of sphericity, *𝓍*^*2*^(435) = 14188.20, *p* = .000, indicated that correlations between items were sufficiently large to conduct an EFA.^36^ Five factors had eigenvalues superior to Kaiser’s^35^ criterion of 1, but the scree plot (see Figure 1) showed inflections that would justify retaining only three factors. Subsequent factor analyses on three-factor varimax and oblimin rotation solutions revealed little difference. Owing to parsimony, interpretability of factors and consistency with the factor structure of the original MPAI-4, a three-factor solution with a varimax rotation method was thus preferred.

**Figure 1.**
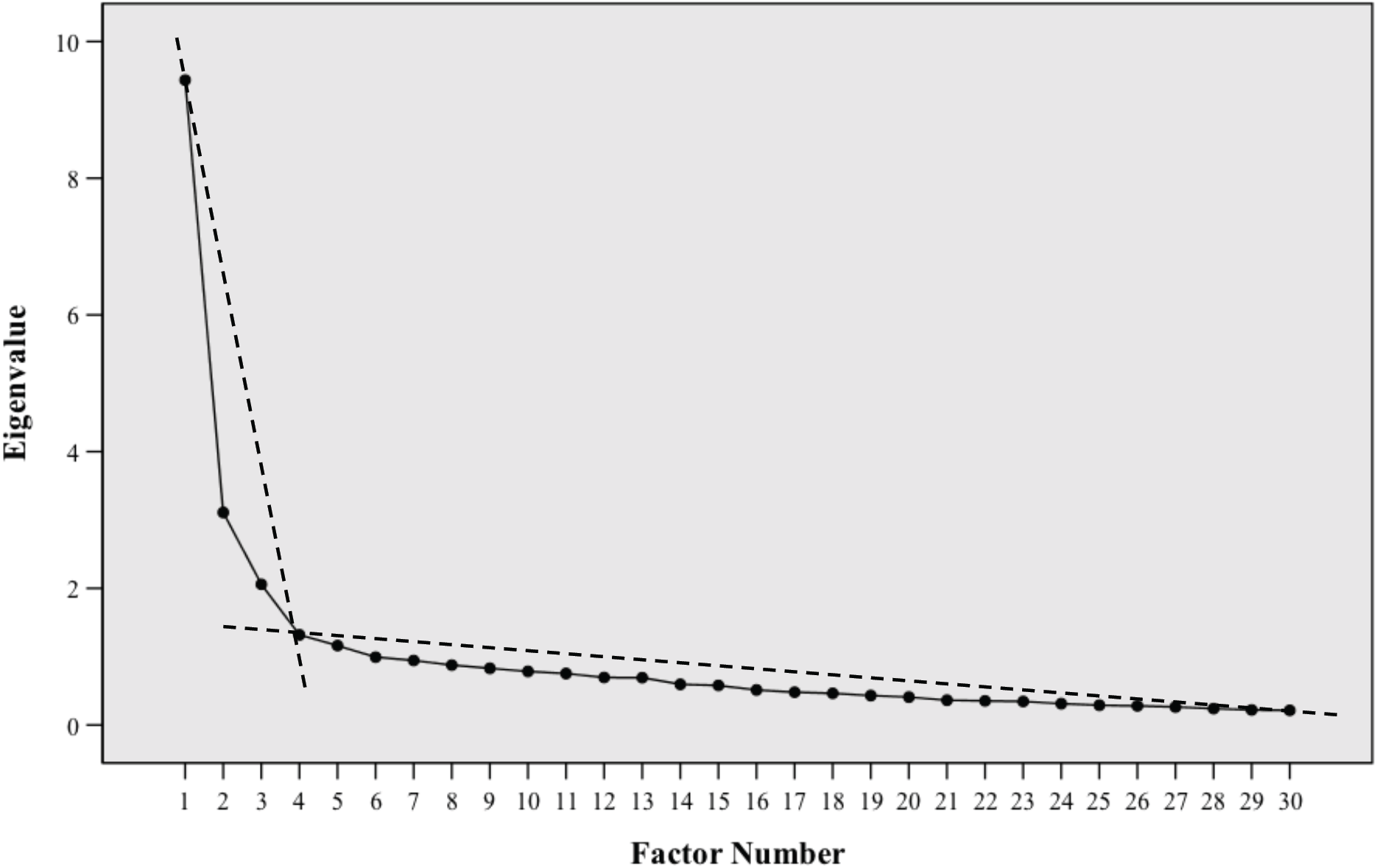
Scree plot obtained by performing an exploratory factor analysis using varimax rotation on z-scores from the 30 items of the French-Canadian MPAI-4.

The final solution accounted for 48.68% of the variance. Communalities, factor loading values and percent of variance for the final solution are presented in Table 2. Items are ordered by their item number to facilitate interpretation. Items allocated to a specific factor were based on loading value *≥* .20 on that factor. A total of 20 items in the final solution also had loading values *≥* .20 on multiple factors, but those items had a higher primary loading value on their assigned factor. Only one of 30 items failed to have a primary factor loading value *≥* .20 on any factor (item 4). Failure to load on a factor reflects the homogeneity of participants’ responses on that specific item, resulting in less variability and positively skewed data. Additional EFAs after eliminating cross-loading items and item 4 did not result in an improved final solution. In order to maintain the questionnaire’s integrity, cross-loading items were retained and item 4 was rationally assigned to the Factor 2.

**Table 2.**
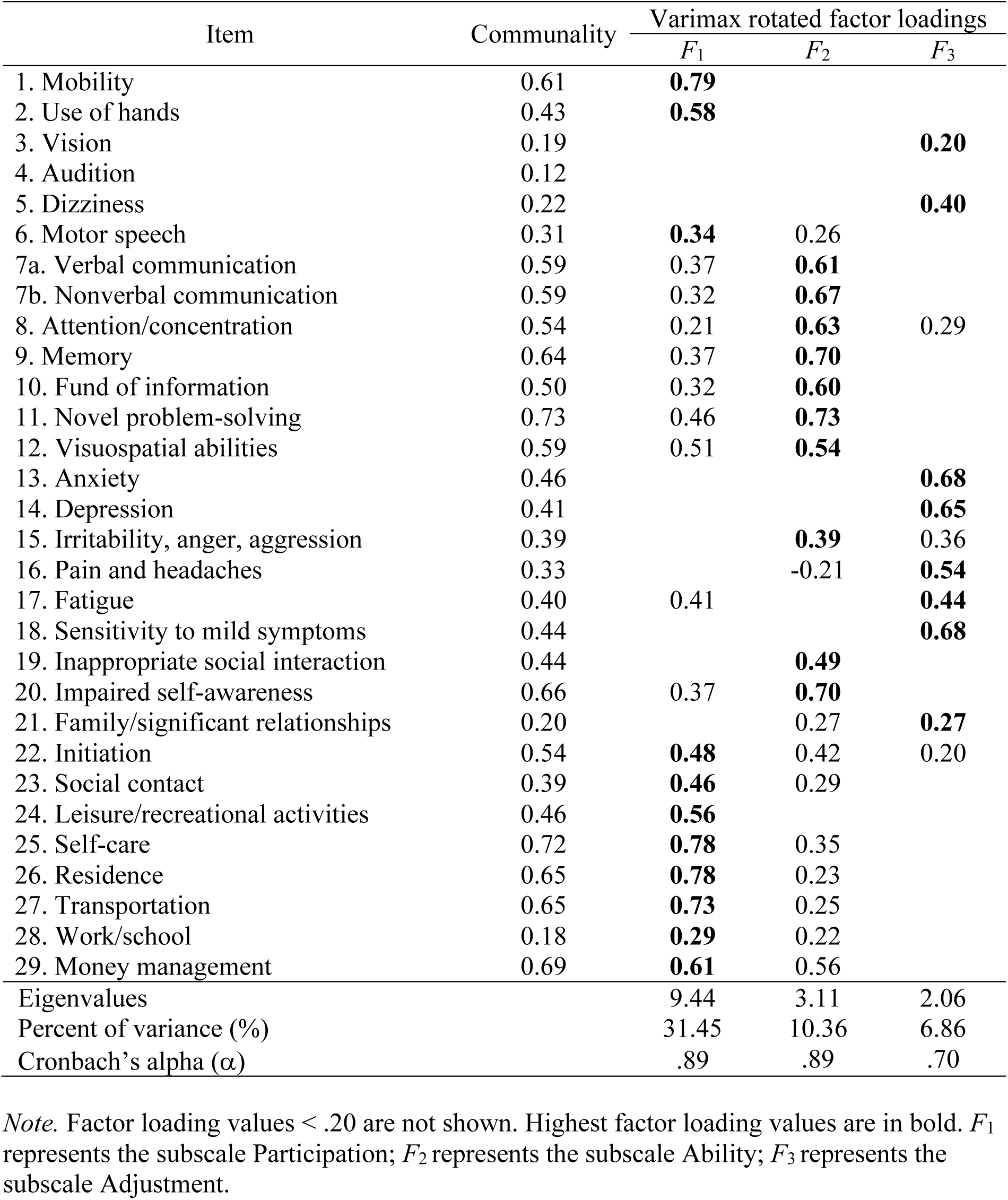
Summary of exploratory factor analysis results for the French-Canadian MPAI-4 questionnaire (N = 1012)

In sum, the three factors extracted using data from the French-Canadian MPAI-4 in a Canadian TBI sample were similar, but not entirely identical to the three subscales found in the original MPAI-4. This may be explained, for example, by sociocultural differences between American and French-Canadian samples, which can affect the distribution of items between factors. However, the factor labels proposed by Malec^6^ also suited the extracted factors and were thus retained. Factor 1, labelled “Participation”, includes 11 items for mobility, motor control, initiative, social contact, independent living, employment status and financial management and accounted for 31.45% of the total variance. Factor 2 labelled “Ability” consists of 10 items, relates to hearing and cognitive abilities, communication and interpersonal interactions and accounted for 10.36% of the total variance. The Factor 3 labelled “Adjustment” accounted for 6.86% of the total variance and consists of 8 items assessing eye vision, symptoms and mood, such as dizziness, anxiety or depression, pain, headaches and fatigue.

### Internal Consistency Reliability Analysis

Values of Cronbach’s alpha for the French-Canadian MPAI-4’s three subscales are presented in Table 2. According to Kline’s^40^ guidelines, the subscales showed good internal consistency, all Cronbach’s alpha values .70 *≤ α ≤* .89. No significant increase in alpha for any of the subscales could have been achieved by eliminating individual items.

### Reference Norms

T score conversion is recommended for clinical use of the MPAI-4, to compare subscale scores and identify areas needing intervention.^41^ Using data from participants included in the validation analyses (*N* = 1012), French-Canadian reference norms for TBI were computed by converting raw scores into T scores. Notably, subscales scores were computed using the original MPAI-4 subscale composition to allow comparison between studies using different versions of the MPAI-4. Conversion tables for the MPAI-4 total and subscales scores can be found online in the Supplementary material (tables S1a to S1d). Tables presenting raw MPAI-4 means and SD according to sex, age, TBI severity, and rehabilitation setting are also included in the Supplementary materials (tables S2a to S2d).

## Discussion

The aim of this paper was to validate the factor structure and establish internal consistency reliability of the French-Canadian MPAI-4 in a Canadian sample of TBI adults receiving post-acute rehabilitation services in a French-speaking setting. We also aimed to provide a set of reference norms to guide clinical use of the MPAI-4. The resulting French-Canadian MPAI-4 was implemented in the clinical practice of four rehabilitation centres in the Greater Montreal region, thus generating MPAI-4 data for all TBI patients receiving post-acute rehabilitation services in those centres. The internal consistency of each subscales in the French-Canadian MPAI-4 was assessed using Cronbach’s alpha and our findings show good internal consistency, with alpha values similar to those reported for the original MPAI-4.^41^ To validate the factor structure of the French-Canadian MPAI-4, we collected all patients’ first MPAI-4 measure and conducted an exploratory factor analysis (EFA). To our knowledge, this study is the first to try and replicate the original MPAI-4 factor structure using another version of the MPAI-4 questionnaire. Our results also show that the factor structure of the French-Canadian MPAI-4 is similar to the original MPAI-4’s, that is, the questionnaire’s items distribute themselves between three factors (i.e. three subscales). However, some discrepancies were found at the item distribution level between the French-Canadian and the original MPAI-4.

First, one item (item 4, audition) did not load on any subscale. To preserve the questionnaire’s integrity, we decided to retain the item 4 and rationally assign it to the Factor 2 “Ability”, as it was attributed to that subscale in the original MPAI-4. We found important and clinical utility in keeping the item 4, as hearing impairment is not reflected in any other item in the questionnaire. Retention of this item also ensures that we maintain the questionnaire’s power of generalization and do not compromise comparison purpose with other studies using different versions of the MPAI-4. Secondly, the composition of the subscales slightly differs between the French-Canadian and the original MPAI-4. The EFA attributed a few items to different subscales when compared to the original MPAI-4. For instance, our results suggest that the items 1 (mobility) and 2 (use of hands) belonged to the Participation subscale, rather than the Ability subscale as in the original MPAI-4.

The small differences in item distribution between subscales can be explained by a few factors, such as sample-specific clinical characteristics and inherent sociocultural differences between American and French-Canadian samples, but also the choice of statistical analysis used to measure construct validity. It is well supported in the literature that different factor structures can be obtained following administration to a second sample due to a combination of sampling variations, sociocultural/linguistic context and applied methodology (for example, see Chen et al.^42^ or Gaskin et al.^43^). Besides those subtle variations in item distribution and in line with Kean et al.’s^16^ conclusions, our results nevertheless show general support for the MPAI-4’s subscale structure developed in previous psychometric studies: the French-Canadian MPAI-4 displays three subscales that represent domains broadly defined as Ability, Adjustment and Participation. Finally, given our EFA results, modifications to the tool (i.e. rearranging item order) so subscales’ score calculation reflected the French-Canadian factor structure was considered. However, such a major change to the questionnaire would rather be detrimental, as comparison purpose with other studies using different versions of the MPAI-4 would no longer be possible.

Lastly, the translation and validation of the French-Canadian MPAI-4 was of primary importance, given the critical lack of available assessment tools for TBI sequelae in French for use in French-Canadian rehabilitation settings. The access to not only a standardized tool, but one that has been translated and validated in the primary language of the setting, allows measuring more accurately TBI patients’ impairments, progress and outcomes. The need to validate the MPAI-4 in a French-Canadian sub-population is also in line with recommendations made by the Center for Disease Control and Prevention^44^ regarding the importance of conducting validation studies of outcome measures among sub-populations in order to expand scientific knowledge of outcomes and establish best practices. Moreover, INESSS-ONF Clinical Practice Guidelines in TBI rehabilitation^21^ also prescribe standardized evaluations of patients’ condition using validated assessment tools in order to orient interventions. The use of the validated French-Canadian MPAI-4 with its specific reference norms will indeed allow professionals to apply best practices, but also guide the use of a common measure between rehabilitation centres, which in turn facilitates clinical discussions and comparisons as well as generates new knowledge regarding TBI outcomes. Overall, the French-Canadian MPAI-4 presents to be a validated TBI assessment tool for use in a French-Canadian post-acute rehabilitation setting. The translation of such a comprehensive and established clinical tool certainly fills a gap in the field of TBI rehabilitation and also adds to the assessment options available for use by French-speaking rehabilitation professionals, individuals with TBI and their significant others.

### Study Limitations

One limitation of this study is the lack of direct comparison of factor structure measures between the original and French-Canadian versions of the MPAI-4. Rasch analyses were used for the development of the original MPAI-4 questionnaire and its factor structure.^41^ In our case, however, it was best to deploy EFA, as our aim was to reveal item distribution and factor structure of the French-Canadian MPAI-4 without a priori manipulations.^45,46^ Consequently, the statistical methodology used does not allow direct comparison and may also lead to a different final solution with slightly different item distribution between factors.

## Conclusions

Using an exploratory factor analysis on data from the French-Canadian version of the MPAI-4 and a Canadian sample of TBI adults receiving rehabilitation services, three factors were extracted and are similar to the three subscales found in the original MPAI-4. Small differences in item distribution across factors can be explained in part by sociocultural and clinical differences between the Canadian and American samples used to establish the factor structure of the questionnaire. The factor labels suggested by Malec^6^ also suited the extracted factors and were thus retained for the French-Canadian MPAI-4 (i.e. Ability, Adjustment, Participation). In sum, the French-Canadian MPAI-4 factor structure is validated, and the questionnaire shows good internal consistency. The French-Canadian MPAI-4, with its reference norms, represents a suitable tool to measure functional evolution, outcomes and social integration of individuals with TBI receiving rehabilitation services in a French-Canadian context.

## Supporting information

ICMJE Form

## Data Availability

Possible availability upon request.

## List of abbreviations

(EFA): Exploratory Factor Analysis
(MPAI-4): Mayo-Portland Adaptability Inventory-4
(TBI): Traumatic Brain Injury.

## Supplementary materials: French-Canadian TBI reference norms and raw scores

**Table S1a.** Conversion of MPAI-4 Total raw scores to T scores (mean = 50; SD = 10) for Canadian TBI adults receiving rehabilitation services in a French-speaking setting (N =1012)

| Raw score | T score | Raw score | T score | Raw score | T score | Raw score | T score |
| --- | --- | --- | --- | --- | --- | --- | --- |
| 0 | N/A | 28 | **42.05** | 56 | **58.39** | 84 | N/A |
| 1 | N/A | 29 | **42.63** | 57 | **58.98** | 85 | **75.32** |
| 2 | N/A | 30 | **43.22** | 58 | **59.56** | 86 | N/A |
| 3 | **27.46** | 31 | **43.80** | 59 | **60.15** | 87 | N/A |
| 4 | N/A | 32 | **44.38** | 60 | **60.73** | 88 | N/A |
| 5 | **28.62** | 33 | **44.97** | 61 | **61.31** | 89 | **77.66** |
| 6 | **29.21** | 34 | **45.55** | 62 | **61.90** | 90 | N/A |
| 7 | **29.79** | 35 | **46.14** | 63 | **62.48** | 91 | **78.82** |
| 8 | **30.38** | 36 | **46.72** | 64 | **63.06** | 92 | N/A |
| 9 | **30.96** | 37 | **47.30** | 65 | **63.65** | 93 | N/A |
| 10 | **31.54** | 38 | **47.89** | 66 | **64.23** | 94 | N/A |
| 11 | **32.13** | 39 | **48.47** | 67 | **64.82** | 95 | N/A |
| 12 | **32.71** | 40 | **49.05** | 68 | **65.40** | 96 | N/A |
| 13 | **33.29** | 41 | **49.64** | 69 | **65.98** | 97 | N/A |
| 14 | **33.88** | 42 | **50.22** | 70 | **66.57** | 98 | N/A |
| 15 | **34.46** | 43 | **50.81** | 71 | **67.15** | 99 | N/A |
| 16 | **35.05** | 44 | **51.39** | 72 | **67.73** | 100 | N/A |
| 17 | **35.63** | 45 | **51.97** | 73 | **68.32** | 101 | N/A |
| 18 | **36.21** | 46 | **52.56** | 74 | **68.90** | 102 | N/A |
| 19 | **36.80** | 47 | **53.14** | 75 | **69.48** | 103 | N/A |
| 20 | **37.38** | 48 | **53.72** | 76 | **70.07** | 104 | N/A |
| 21 | **37.96** | 49 | **54.31** | 77 | **70.65** | 105 | N/A |
| 22 | **38.55** | 50 | **54.89** | 78 | **71.24** | 106 | N/A |
| 23 | **39.13** | 51 | **55.48** | 79 | **71.82** | 107 | N/A |
| 24 | **39.71** | 52 | **56.06** | 80 | **72.40** | 108 | N/A |
| 25 | **40.30** | 53 | **56.64** | 81 | **72.99** | 109 | N/A |
| 26 | **40.88** | 54 | **57.23** | 82 | N/A | 110 | N/A |
| 27 | **41.47** | 55 | **57.81** | 83 | **74.15** | 111 | N/A |

*Note.* N/A T score = Raw score not obtained within the sample. According to Malec and Lezak<sup>41</sup>: T scores between 40 and 60 would be considered average or typical of people involved in outpatient rehabilitation following acquired brain injury (ABI); T scores between 40 and 50 may be considered in the mild to moderate range of overall severity compared to other people with ABI; T scores between 50 and 60, may be considered in the moderate to severe range compared to other people with ABI. Hence, T scores above 60 would suggest severe limitations even as compared to other people with ABI, T scores between 30 and 40 suggest mild limitations, and T scores below 30 represent relatively good outcomes.

**Table S1b.** Conversion of MPAI-4 Ability subscale raw scores to T scores (mean = 50; SD = 10) for Canadian TBI adults receiving rehabilitation services in a French-speaking setting (N =1012)

| Raw score | T score | Raw score | T score | Raw score | T score |
| --- | --- | --- | --- | --- | --- |
| 0 | **32.34** | 16 | **53.26** | 32 | **74.19** |
| 1 | **33.65** | 17 | **54.57** | 33 | **75.49** |
| 2 | **34.96** | 18 | **55.88** | 34 | **76.80** |
| 3 | **36.26** | 19 | **57.19** | 35 | **78.11** |
| 4 | **37.57** | 20 | **58.49** | 36 | **79.42** |
| 5 | **38.88** | 21 | **59.80** | 37 | **N/A** |
| 6 | **40.19** | 22 | **61.11** | 38 | **82.03** |
| 7 | **41.50** | 23 | **62.42** | 39 | **83.34** |
| 8 | **42.80** | 24 | **63.73** | 40 | **N/A** |
| 9 | **44.11** | 25 | **65.03** | 41 | **N/A** |
| 10 | **45.42** | 26 | **66.34** | 42 | **N/A** |
| 11 | **46.73** | 27 | **67.65** | 43 | **N/A** |
| 12 | **48.03** | 28 | **68.96** | 44 | **N/A** |
| 13 | **49.34** | 29 | **70.26** | 45 | **N/A** |
| 14 | **50.65** | 30 | **71.57** | 46 | **N/A** |
| 15 | **51.96** | 31 | **72.88** | 47 | **N/A** |

*Note.* N/A T score = Raw score not obtained within the sample. According to Malec and Lezak<sup>41</sup>: T scores between 40 and 60 would be considered average or typical of people involved in outpatient rehabilitation following acquired brain injury (ABI); T scores between 40 and 50 may be considered in the mild to moderate range of overall severity compared to other people with ABI; T scores between 50 and 60, may be considered in the moderate to severe range compared to other people with ABI. Hence, T scores above 60 would suggest severe limitations even as compared to other people with ABI, T scores between 30 and 40 suggest mild limitations, and T scores below 30 represent relatively good outcomes.

**Table S1c.** Conversion of MPAI-4 Adjustment subscale raw scores to T scores (mean = 50; SD = 10) for Canadian TBI adults receiving rehabilitation services in a French-speaking setting (N =1012)

| Raw score | T score | Raw score | T score | Raw score | T score |
| --- | --- | --- | --- | --- | --- |
| 0 | **23.01** | 16 | **46.73** | 32 | **70.46** |
| 1 | **24.50** | 17 | **48.22** | 33 | **71.94** |
| 2 | **25.98** | 18 | **49.70** | 34 | **73.42** |
| 3 | **27.46** | 19 | **51.18** | 35 | **74.90** |
| 4 | **28.94** | 20 | **52.67** | 36 | **76.39** |
| 5 | **30.43** | 21 | **54.15** | 37 | **77.87** |
| 6 | **31.91** | 22 | **55.63** | 38 | **79.35** |
| 7 | **33.39** | 23 | **57.11** | 39 | **N/A** |
| 8 | **34.87** | 24 | **58.60** | 40 | **N/A** |
| 9 | **36.36** | 25 | **60.08** | 41 | **N/A** |
| 10 | **37.84** | 26 | **61.56** | 42 | **N/A** |
| 11 | **39.32** | 27 | **63.04** | 43 | **N/A** |
| 12 | **40.80** | 28 | **64.53** | 44 | **N/A** |
| 13 | **42.29** | 29 | **66.01** | 45 | **N/A** |
| 14 | **43.77** | 30 | **67.49** | 46 | **N/A** |
| 15 | **45.25** | 31 | **68.97** |  |  |

*Note.* N/A T score = Raw score not obtained within the sample. According to Malec and Lezak<sup>41</sup>: T scores between 40 and 60 would be considered average or typical of people involved in outpatient rehabilitation following acquired brain injury (ABI); T scores between 40 and 50 may be considered in the mild to moderate range of overall severity compared to other people with ABI; T scores between 50 and 60, may be considered in the moderate to severe range compared to other people with ABI. Hence, T scores above 60 would suggest severe limitations even as compared to other people with ABI, T scores between 30 and 40 suggest mild limitations, and T scores below 30 represent relatively good outcomes.

**Table S1d.** Conversion of MPAI-4 Participation subscale raw scores to T scores (mean = 50; SD = 10) for Canadian TBI adults receiving rehabilitation services in a French-speaking setting (N =1012)

| Raw score | T score | Raw score | T score |
| --- | --- | --- | --- |
| 0 | **27.78** | 16 | **49.07** |
| 1 | **29.12** | 17 | **50.40** |
| 2 | **30.45** | 18 | **51.74** |
| 3 | **31.78** | 19 | **53.07** |
| 4 | **33.11** | 20 | **54.40** |
| 5 | **34.44** | 21 | **55.73** |
| 6 | **35.77** | 22 | **57.06** |
| 7 | **37.10** | 23 | **58.39** |
| 8 | **38.43** | 24 | **59.72** |
| 9 | **39.76** | 25 | **61.05** |
| 10 | **41.09** | 26 | **62.38** |
| 11 | **42.42** | 27 | **63.71** |
| 12 | **43.75** | 28 | **65.04** |
| 13 | **45.08** | 29 | **66.37** |
| 14 | **46.41** | 30 | **67.70** |
| 15 | **47.74** |  |  |

*Note.* According to Malec and Lezak<sup>41</sup>: T scores between 40 and 60 would be considered average or typical of people involved in outpatient rehabilitation following acquired brain injury (ABI); T scores between 40 and 50 may be considered in the mild to moderate range of overall severity compared to other people with ABI; T scores between 50 and 60, may be considered in the moderate to severe range compared to other people with ABI. Hence, T scores above 60 would suggest severe limitations even as compared to other people with ABI, T scores between 30 and 40 suggest mild limitations, and T scores below 30 represent relatively good outcomes.

**Table S2a.** Descriptive raw data for the MPAI-4 according to sex, for a sample of Canadian TBI adults receiving rehabilitation services in a French-speaking setting (N =1011)

|  | Mean (SD) |
| --- | --- |
| **Male (N = 625)** |  |
| Ability subscale | 13.99 (7.78) |
| Adjustment subscale | 17.91 (6.88) |
| Participation subscale | 17.03 (7.41) |
| MPAI-4 total score | 42.11 (17.39) |
| **Female (N = 386)** |  |
| Ability subscale | 12.66 (7.32) |
| Adjustment subscale | 18.62 (6.49) |
| Participation subscale | 16.13 (7.62) |
| MPAI-4 total score | 40.72 (16.67) |

**Table S2b.** Descriptive raw data for the MPAI-4 according to age, for a sample of Canadian TBI adults receiving rehabilitation services in a French-speaking setting (N =1012)

|  | Mean (SD) |
| --- | --- |
| **Ages 14-19 years (N = 30)** |  |
| Ability subscale | 11.47 (7.44) |
| Adjustment subscale | 17.37 (7.05) |
| Participation subscale | 14.47 (7.30) |
| MP AI-4 total score | 36.83 (16.93) |
| **Ages 20-29 years (N = 127)** |  |
| Ability subscale | 12.48 (8.62) |
| Adjustment subscale | 18.46 (6.73) |
| Participation subscale | 15.23 (7.80) |
| MP AI-4 total score | 39.53 (18.30) |
| **Ages 30-39 years (N = 117)** |  |
| Ability subscale | 10.74 (6.97) |
| Adjustment subscale | 18.20 (7.23) |
| Participation subscale | 13.66 (6.85) |
| MP AI-4 total score | 36.91 (16.04) |
| **Ages 40-49 years (N = 111)** |  |
| Ability subscale | 12.30 (7.38) |
| Adjustment subscale | 19.41 (6.43) |
| Participation subscale | 15.04 (6.81) |
| MP AI-4 total score | 40.15 (15.76) |
| **Ages 50-59 years (N = 159)** |  |
| Ability subscale | 12.67 (7.49) |
| Adjustment subscale | 18.08 (6.12) |
| Participation subscale | 14.85 (7.14) |
| MP AI-4 total score | 39.34 (16.50) |
| **Ages 60-69 years (N = 146)** |  |
| Ability subscale | 13.57 (7.00) |
| Adjustment subscale | 17.63 (6.87) |
| Participation subscale | 16.48 (7.30) |
| MP AI-4 total score | 40.83 (16.90) |
| **Ages 70-79 years (N = 168)** |  |
| Ability subscale | 15.36 (7.50) |
| Adjustment subscale | 17.94 (7.46) |
| Participation subscale | 19.27 (7.26) |
| MP AI-4 total score | 45.25 (18.02) |
| **Ages 80 years and above (N = 154)** |  |
| Ability subscale | 16.37 (7.01) |
| Adjustment subscale | 18.21 (6.24) |
| Participation subscale | 21.09 (6.29) |
| MP AI-4 total score | 48.05 (15.39) |

**Table S2c.** Descriptive raw data for the MPAI-4 according to TBI severity, for a sample of Canadian TBI adults receiving rehabilitation services in a French-speaking setting (N =1012)

|  | Mean (SD) |
| --- | --- |
| Mild (N = 229) |  |
| Ability subscale | 8.68 (4.88) |
| Adjustment subscale | 18.11 (6.26) |
| Participation subscale | 10.61 (5.09) |
| MPAI-4 total score | 32.04 (12.18) |
| Complicated Mild (N = 229) |  |
| Ability subscale | 14.38 (7.67) |
| Adjustment subscale | 17.68 (6.93) |
| Participation subscale | 18.68 (7.34) |
| MPAI-4 total score | 43.55 (17.36) |
| Moderate (N = 371) |  |
| Ability subscale | 13.45 (6.94) |
| Adjustment subscale | 17.36 (6.50) |
| Participation subscale | 17.45 (6.86) |
| MPAI-4 total score | 41.35 (16.07) |
| Severe (N = 183) |  |
| Ability subscale | 18.48 (8.16) |
| Adjustment subscale | 20.65 (7.08) |
| Participation subscale | 20.40 (7.10) |
| MPAI-4 total score | 51.96 (17.73) |

**Table S2d.**
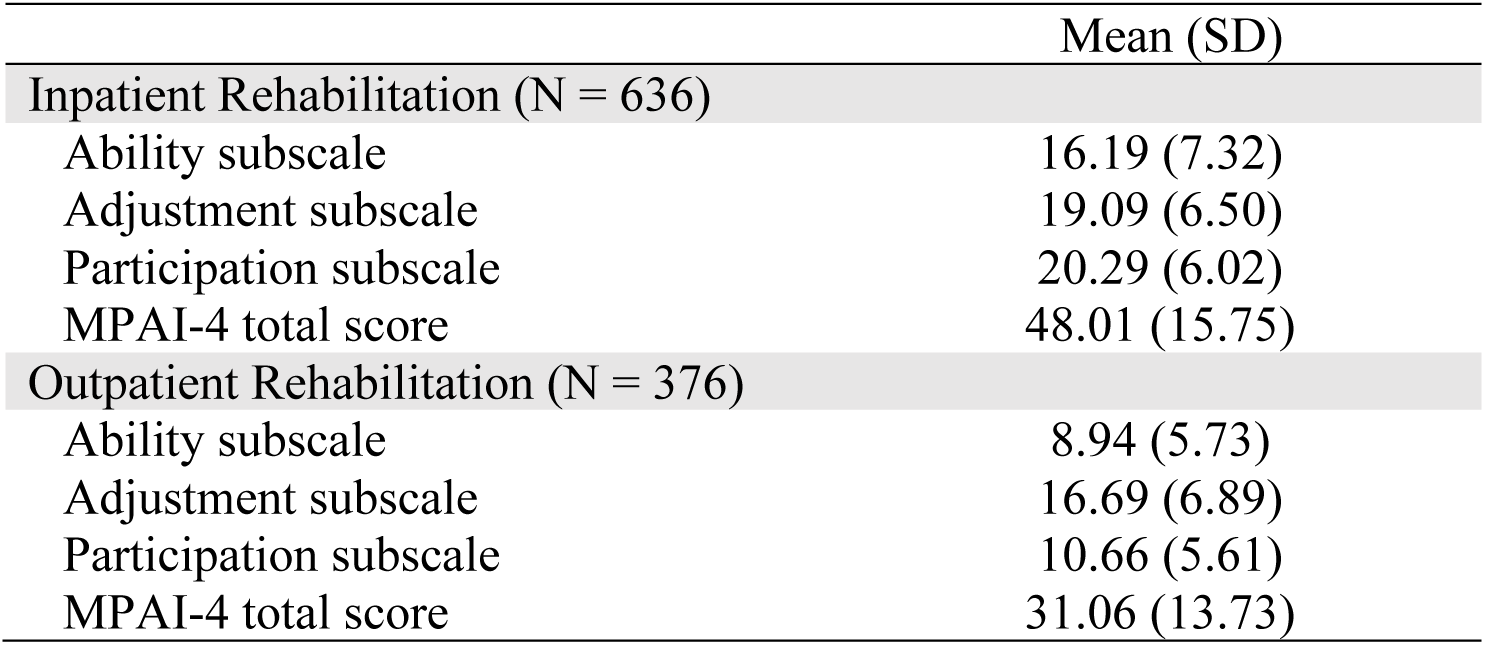
Descriptive raw data for the MPAI-4 according to rehabilitation setting, for a sample of Canadian TBI adults receiving rehabilitation services in a French-speaking setting (N =1012)

## Author Guidelines for Reporting Scale Development and Validation Results

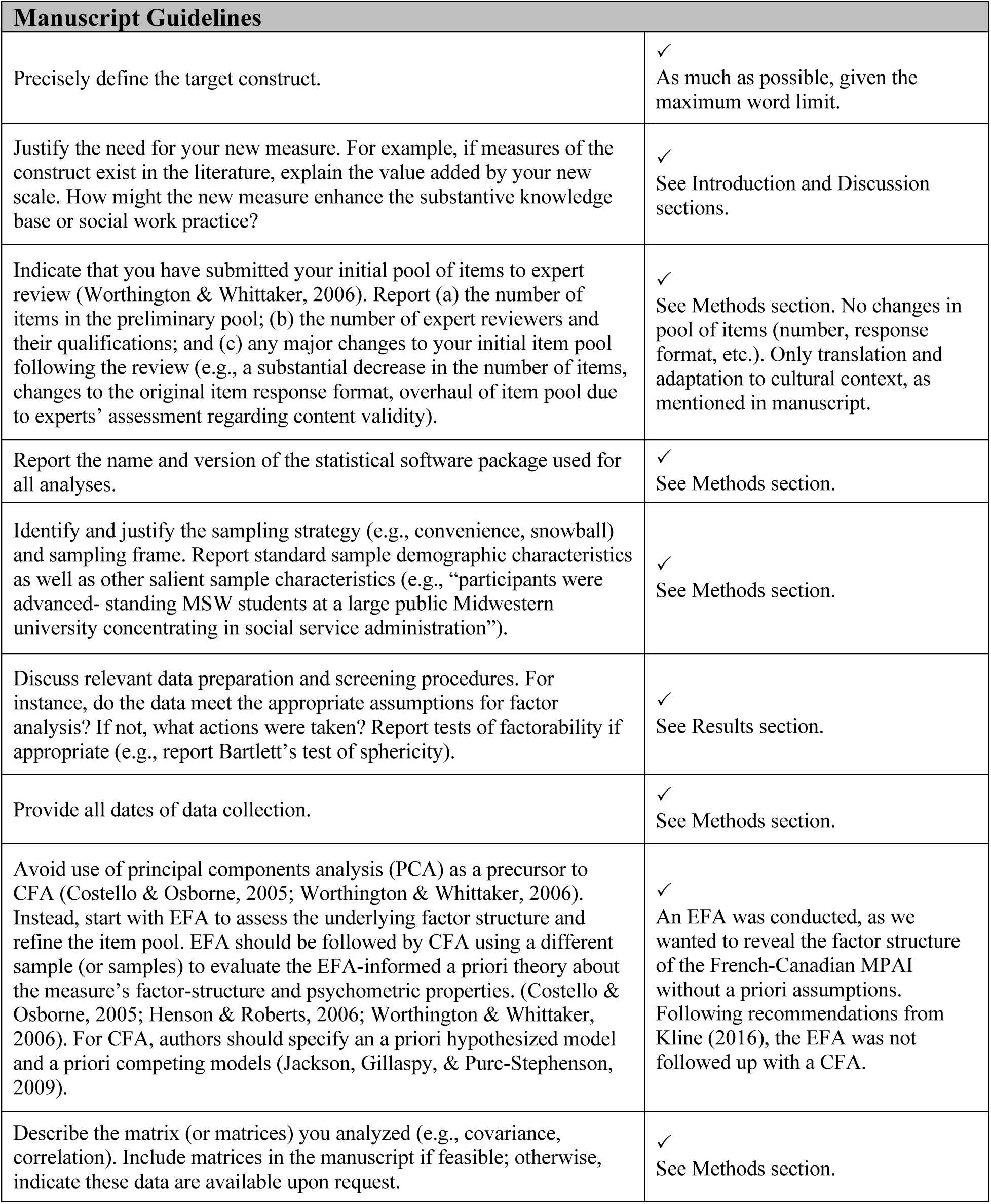
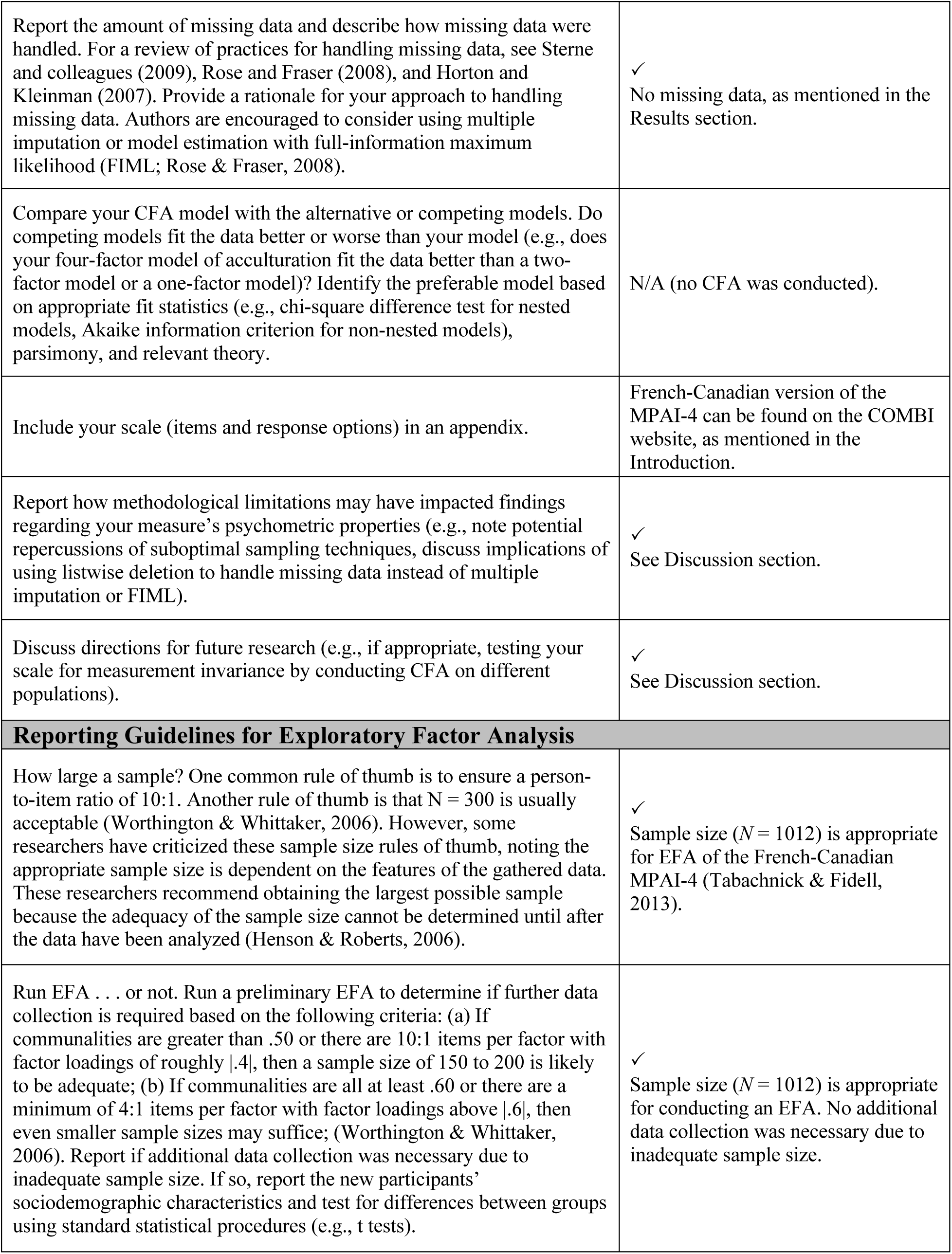
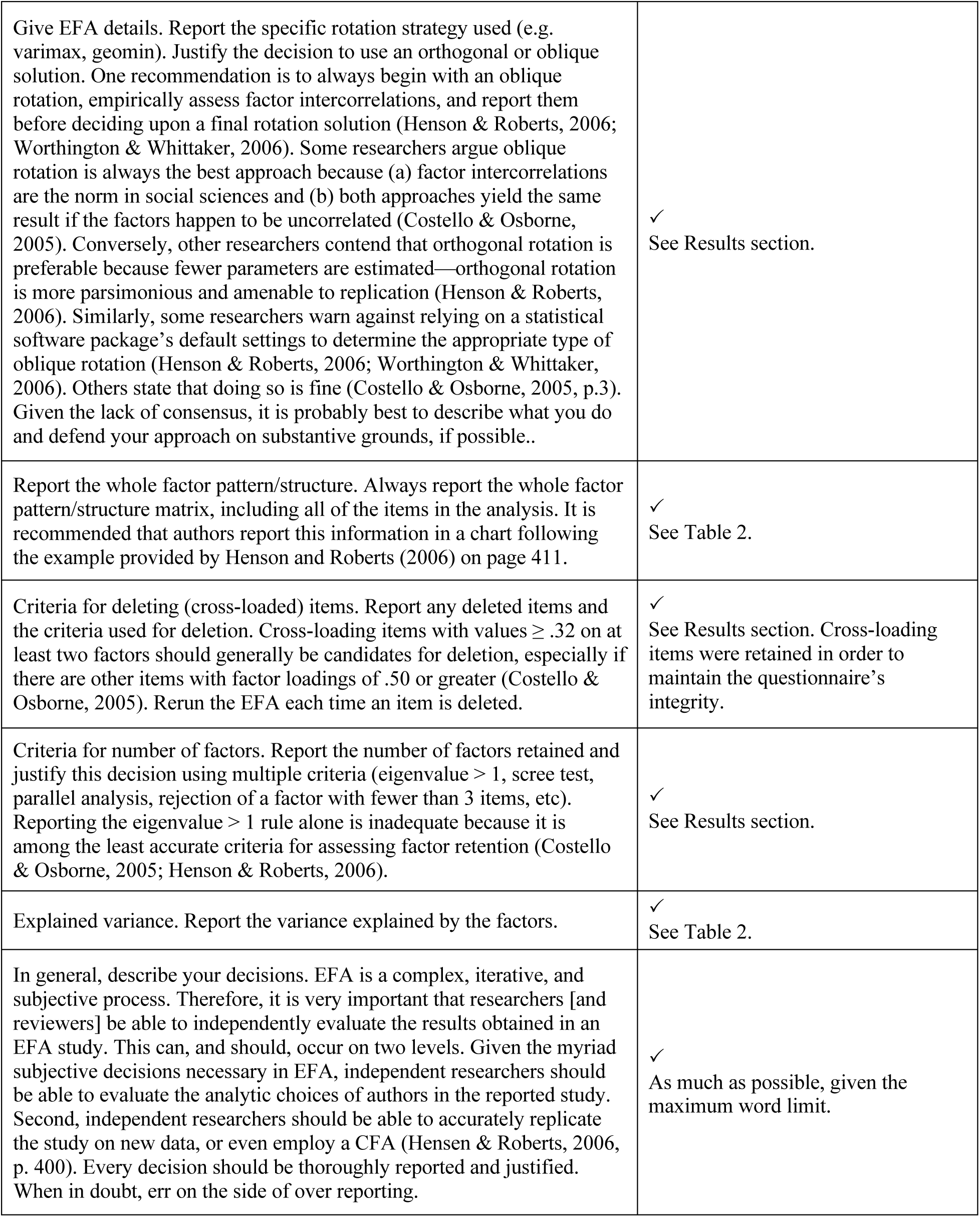

